# Frequency and antibiotic susceptibility of urinary *Klebsiell*a in Lahore

**DOI:** 10.64898/2026.11.15.26368163

**Authors:** Zunaira Kalsoom, Muhammad Mohsin Khan, Syeda Rija Rashid

## Abstract

Urinary tract infections are commonly caused by bacteria whose antibiotic susceptibility varies between clinical settings.^1^ We studied urinary isolates from 95 patients with suspected infection at Lahore General Hospital, Pakistan, between July and December 2023. Urine culture showed significant bacterial growth in 63 specimens (66.3%). *Escherichia coli* accounted for 30 isolates, followed by 18 *Klebsiella* isolates, comprising 16 *Klebsiella pneumoniae* and two *Klebsiella oxytoca. Klebsiella* was recovered from 12 female and six male patients, and the largest number of *Klebsiella* isolates occurred in patients aged 21–30 years. The aggregate susceptibility results, recorded for 18 isolates per antibiotic, showed the highest susceptibility to imipenem and meropenem (17/18 each), followed by nitrofurantoin (16/18). Resistance to several cephalosporins, fluoroquinolones and aminoglycosides was frequent. These findings describe the bacterial distribution and reported susceptibility patterns in a small hospital sample. Larger studies with verified species-specific susceptibility data are needed before these findings can guide local treatment policy.

## Introduction

Urinary tract infections affect the bladder, urethra and kidneys and are encountered in both community and hospital practice. Their clinical presentation ranges from lower urinary symptoms to systemic infection. Diagnosis requires consideration of symptoms, patient characteristics and laboratory findings.^2^ Differences in underlying illness and infection severity also influence how susceptibility data should be interpreted across patient groups.^3^

*Klebsiella pneumoniae* is an important urinary pathogen, particularly in patients with underlying illness or impaired host defences.^4^ Its capsule, adhesins, iron-acquisition systems and capacity to form biofilms contribute to colonisation and persistence.^5^ Extended-spectrum β-lactamases and carbapenemases can limit the antibiotics available for treatment.^6,7^ These mechanisms cannot, however, be inferred from a susceptibility percentage alone.

Studies from South Punjab, Timurgara, Karachi and Quetta have described antibiotic resistance among urinary pathogens in Pakistan.^8,9,10,11^ Their findings support the use of local laboratory data, because the distribution of organisms and their susceptibility patterns vary between hospitals and patient populations. Studies of *Klebsiella* elsewhere have also documented variation in resistance, virulence characteristics and genetic diversity.^12,13^

This study examined urine specimens from patients with suspected urinary tract infection at Lahore General Hospital. The objectives were to describe the frequency of *Klebsiella* among recovered organisms and summarise the reported antibiotic susceptibility of the isolates.

## Methods

A cross-sectional study was conducted at Lahore General Hospital, Lahore, Pakistan, from July to December 2023. Patients with suspected urinary tract infection were recruited using non-probability convenience sampling. Both sexes and all age groups were eligible. The study included 95 patients. The sample size was based on an anticipated proportion of 0.14, a 95% confidence level and an absolute precision of 0.07, using n = Z^2^P(1 − P)/d^2^. The planned sample size was 95. Patients without suspected infection, patients with the chronic diseases listed in the study protocol, and improperly labelled or contaminated specimens were excluded.

Urine specimens were collected in sterile screw-capped containers after patients were instructed to clean the external genital area. Specimens were transported to the microbiology laboratory within one hour. Samples awaiting examination were refrigerated at 4–8 °C. Urine was inoculated onto cysteine-lactose-electrolyte-deficient agar and incubated aerobically overnight at 37 °C. Cultures were assessed for significant bacterial growth. Cultured organisms were examined by Gram staining and biochemical testing. The described tests included urease production, citrate utilisation, triple sugar iron agar reactions, indole production, motility and oxidase activity. An API 20E identification system was also described. Inoculated API strips were incubated at 37 °C for 24 hours, and reaction codes were compared with the manufacturer’s identification catalogue.

Susceptibility was assessed by the Kirby–Bauer disc diffusion method on Mueller–Hinton agar (Hi-Media). Three to five colonies were suspended in sterile saline and adjusted to a turbidity standard. The suspension was spread across the agar surface, antibiotic discs were applied, and plates were incubated aerobically overnight at 35 °C. Inhibition-zone diameters were measured with a calibrated scale and interpreted using a zone-interpretation chart. The tested agents and recorded susceptible and resistant counts are listed in Table 3. Data were summarised as frequencies and percentages using SPSS version 20.0. Culture outcomes were expressed as proportions of the 95 sampled patients, and organism frequencies as proportions of the 63 recovered isolates. The proportion of each Klebsiella species used the 18 Klebsiella isolates as the denominator. Sex-specific Klebsiella isolation proportions used the total number of sampled patients of each sex. Susceptibility percentages used the 18 recorded results per antibiotic. Percentages in the tables and figures were calculated from the reported counts and rounded to one decimal place. No inferential tests or treatment-outcome analyses were reported.

## Results

### Patients and culture outcomes

The study included 95 patients: 55 females (57.9%) and 40 males (42.1%). Significant bacterial growth was reported in 63 specimens (66.3%); the remaining 32 specimens (33.7%) were culture negative (Table 1). These percentages describe the recruited hospital sample and do not estimate the prevalence of urinary tract infection in the wider population.

**Table 1.** Patient characteristics and urine culture outcomes.

| **Characteristic** | **Category** | **n/N** | **Percentage (%)** |
|---|---|---|---|
| Sex | Female | 55/95 | 57.9 |
| Sex | Male | 40/95 | 42.1 |
| Urine culture | Positive | 63/95 | 66.3 |
| Urine culture | Negative | 32/95 | 33.7 |
| *Klebsiella* isolation | All patients | 18/95 | 18.9 |
| *Klebsiella* isolation | Female patients | 12/55 | 21.8 |
| *Klebsiella* isolation | Male patients | 6/40 | 15.0 |

N = 95 patients. Percentages use the denominator shown in each row. Sex-specific *Klebsiella* percentages are descriptive and were calculated from the reported counts; no significance test is presented.

### Distribution of bacterial isolates

*Escherichia coli* was the most frequently recovered organism, with 30 isolates (47.6% of the 63 isolates). *Klebsiella* accounted for 18 isolates (28.6%), followed by *Staphylococcus* species (7; 11.1%), *Enterococcus* (5; 7.9%), *Pseudomonas aeruginosa* (2; 3.2%) and *Proteus mirabilis* (1; 1.6%) (Table 2 and Fig. 1). Gram-negative organisms accounted for 51 isolates (81.0%) and Gram-positive organisms for 12 (19.0%).

**Table 2.** Distribution of bacteria recovered from urine.

| **Bacterial group** | **Isolates (n)** | **Percentage (%)** |
|---|---|---|
| *Escherichia coli* | 30 | 47.6 |
| *Klebsiella* species | 18 | 28.6 |
| *Staphylococcus* species | 7 | 11.1 |
| *Enterococcus* | 5 | 7.9 |
| *Pseudomonas aeruginosa* | 2 | 3.2 |
| *Proteus mirabilis* | 1 | 1.6 |
| Total | 63 | 100.0 |

Denominator = 63 isolates. The 18 *Klebsiella* isolates comprised 16 *K. pneumoniae* (88.9%) and two *K. oxytoca* (11.1%). Percentages were calculated from the reported counts and rounded to one decimal place. Gram-negative and Gram-positive organisms accounted for 51 and 12 isolates, respectively.

**Fig. 1.**
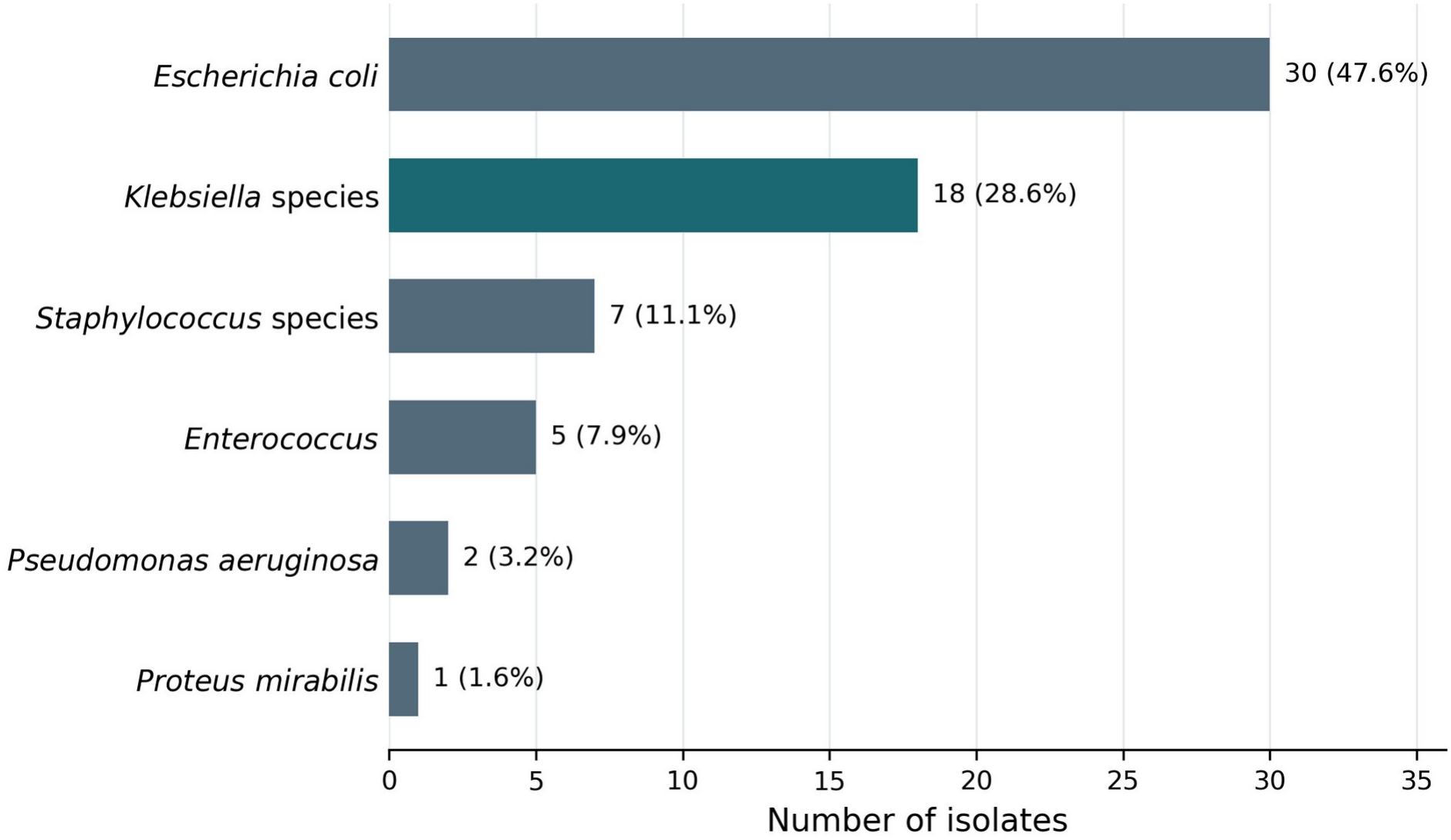
Bacterial distribution among positive urine cultures. Bars show the number of isolates in each bacterial group; labels give counts and percentages of the 63 isolates. *Klebsiella* species include 16 *K. pneumoniae* and two *K. oxytoca*. The figure was prepared from the counts reported in Table 2.

Of the 18 *Klebsiella* isolates, 16 (88.9%) were identified as *K. pneumoniae* and two (11.1%) as *K. oxytoca. Klebsiella* was recovered from 12 female and six male patients, corresponding to 21.8% of the 55 females and 15.0% of the 40 males. These were descriptive comparisons; no test of a sex-associated difference was reported.

The largest number of *Klebsiella* isolates occurred in patients aged 21–30 years (8/18; 44.4%), followed by those aged 31–40 years (4/18; 22.2%). Two isolates each were reported in the 11–20 and 41–50 year groups, and one each in the youngest reported group and the 51–60 year group. Age-specific infection risk could not be estimated because the numbers of all recruited patients in each age group were not available.

### Reported antibiotic susceptibility

The susceptibility results contained 18 observations for each antibiotic (Table 3 and Fig. 2). Imipenem and meropenem each had 17 susceptible isolates (94.4%), followed by nitrofurantoin with 16 (88.9%) and fosfomycin with 14 (77.8%). Amikacin had four susceptible isolates (22.2%), while tobramycin and amoxicillin–clavulanic acid each had three (16.7%).

**Table 3.** Reported antibiotic susceptibility of urinary isolates.

| **Antibiotic** | **Susceptible, n (%)** | **Resistant, n (%)** |
|---|---|---|
| Nitrofurantoin | 16 (88.9) | 2 (11.1) |
| Fosfomycin | 14 (77.8) | 4 (22.2) |
| Amikacin | 4 (22.2) | 14 (77.8) |
| Tobramycin | 3 (16.7) | 15 (83.3) |
| Gentamicin | 2 (11.1) | 16 (88.9) |
| Ciprofloxacin | 2 (11.1) | 16 (88.9) |
| Levofloxacin | 2 (11.1) | 16 (88.9) |
| Imipenem | 17 (94.4) | 1 (5.6) |
| Meropenem | 17 (94.4) | 1 (5.6) |
| Cefotaxime | 2 (11.1) | 16 (88.9) |
| Ceftazidime | 2 (11.1) | 16 (88.9) |
| Tigecycline | 2 (11.1) | 16 (88.9) |
| Tazobactam (TZP)† | 1 (5.6) | 17 (94.4) |
| Amoxicillin–clavulanic acid | 3 (16.7) | 15 (83.3) |

Denominator = 18 recorded results for each antibiotic. Percentages were calculated from counts and rounded to one decimal place. Results are aggregate *Klebsiella* data; species-specific results are unavailable. Intermediate counts were not reported. †The source lists “Tazobactum (TZP)”; the tested formulation and disc code require confirmation. The recorded susceptibility categories require verification against the laboratory standard used.

**Fig. 2.**
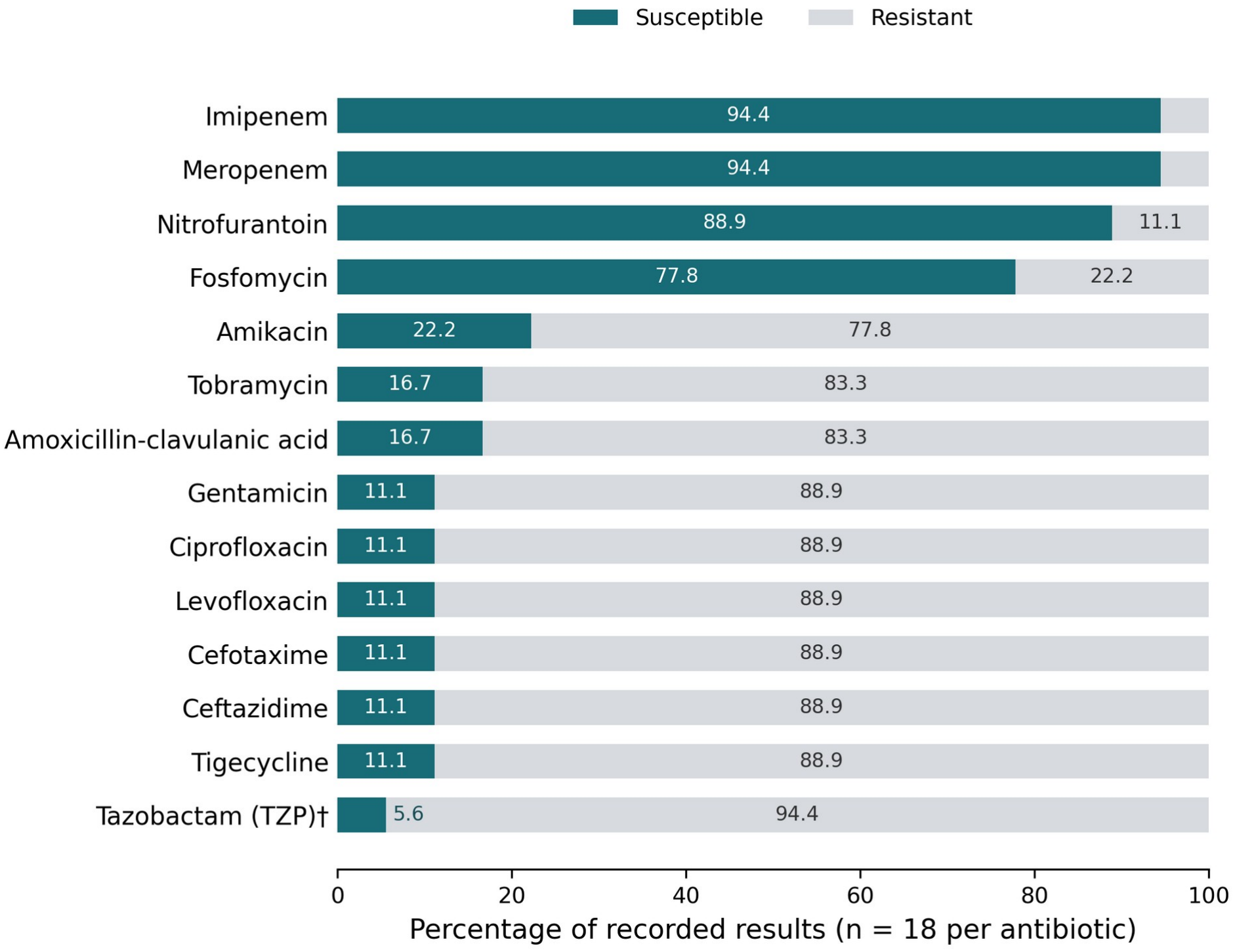
Reported antibiotic susceptibility of urinary isolates. Bars show the percentages classified as susceptible or resistant among 18 recorded results per antibiotic. Drugs are ordered by the number classified as susceptible. Data are shown in Table 3. Species-specific results and intermediate counts were not available. †The source label “Tazobactam (TZP)” requires confirmation. These descriptive results do not establish clinical treatment efficacy.

Gentamicin, ciprofloxacin, levofloxacin, cefotaxime, ceftazidime and tigecycline each had two susceptible isolates (11.1%) and 16 resistant isolates (88.9%). The agent recorded as tazobactam (TZP) had one susceptible isolate and 17 resistant isolates. Its exact antibiotic formulation requires confirmation. The aggregate results do not separate *K. pneumoniae* from *K. oxytoca*.

## Discussion

*Klebsiella* was the second most frequently recovered bacterial group after *E. coli* in this hospital sample. Culture positivity was 66.3%, while *Klebsiella* accounted for 28.6% of recovered isolates and 18.9% of all sampled patients. These denominators answer different questions and should not be used interchangeably. Reports from Pakistan and other settings also identify *E. coli* and *Klebsiella* among urinary pathogens, although their relative frequencies vary.^8,9,10,14,15^

The high reported susceptibility to imipenem and meropenem is consistent with some earlier studies of urinary *Klebsiella* isolates. Polse and colleagues reported susceptibility to both agents, and studies from Iraq and India also described activity of imipenem against urinary isolates.^15,16,17^ However, those studies differed in patient selection, collection period and testing procedures.

The present results should therefore be regarded as a local descriptive observation rather than a basis for recommending carbapenems for empirical treatment.

Nitrofurantoin and fosfomycin also showed relatively high reported susceptibility in this sample. Other studies have described variation in the activity of urinary antibiotics between organisms and clinical settings.^18,19^ A laboratory susceptibility result does not by itself establish suitability for every urinary syndrome. Patient characteristics, the site and severity of infection, and validated interpretation of the laboratory result remain necessary when selecting treatment.^3^ Resistance to the tested cephalosporins, fluoroquinolones and several aminoglycosides was frequent. This pattern is consistent with the concern about restricted treatment options described in studies of resistant *Klebsiella* infections.^12,13,20^ The study did not include molecular testing or a reported confirmatory test for extended-spectrum β-lactamase or carbapenemase production. Specific resistance mechanisms therefore cannot be assigned to these isolates.

Studies of hypervirulent and invasive *K. pneumoniae* have shown that clinical presentation, underlying illness and bacterial characteristics are relevant to interpretation.^21,22,23^ None of those findings establishes that the isolates in the present study were hypervirulent. The available data describe culture identification and aggregate susceptibility; they do not describe virulence genes, transmission, treatment response or mortality.

Species identification also matters. Two of the 18 *Klebsiella* isolates were reported as *K. oxytoca*, and the susceptibility results could not be separated by species. Studies comparing conventional and genotypic identification methods illustrate the need to confirm organism identification when interpreting *Klebsiella* data.^24^ Future surveillance should retain isolate-level species identification alongside each antibiotic result.

More *Klebsiella* isolates were recovered from females than males, but females also made up a larger proportion of the recruited patients. The relevant descriptive proportions were 12/55 and 6/40, rather than the shares of females and males among the 18 positive patients. The findings do not establish a sex-related risk difference. Broader guidance on recurrent infection in women likewise depends on clinical history and the population being assessed, rather than isolate counts alone.^25^

## Limitations

This was a small, single-hospital study using convenience sampling. Only 18 *Klebsiella* isolates were reported, so a change in the result for one isolate changes an antibiotic percentage by approximately 5.6 percentage points. The aggregate data did not allow species-specific susceptibility estimates or assessment of multidrug resistance in individual isolates. Age-group denominators, molecular resistance data and treatment outcomes were unavailable. Patient-reported symptoms, medication history and previous antibiotic use could also have introduced reporting error. The susceptibility categories require verification against the laboratory’s original antibiotic-specific interpretive criteria.

## Conclusion

*E. coli* was the most frequently recovered organism, followed by *Klebsiella*, in urine specimens from patients at Lahore General Hospital. The reported aggregate *Klebsiella* results showed higher susceptibility to imipenem, meropenem, nitrofurantoin and fosfomycin than to several other tested agents. These findings support continued local surveillance, with clear species identification, validated susceptibility interpretation and linkage to clinical outcomes.

## Statements

### Ethics approval and informed consent

Ethical clearance was obtained from an ethical review board. Permission was obtained from the Institute of Public Health, Lahore, and the study setting. Patient consent was obtained before specimen collection and data recording.

### Conflict of interest

The authors declare no competing interests.

### Author contributions

Zunaira Kalsoom conceived the study, carried out the laboratory work, collected and analysed the data, and drafted the manuscript. Muhammad Mohsin Khan reviewed the analysis and reviewed and edited the manuscript for content and language. Syeda Rija Rashid contributed to the methodology, including sampling and susceptibility testing. All authors read and approved the final manuscript.

### Funding

This research received no specific grant from any funding agency in the public, commercial or not-for-profit sectors.

### Data availability

The data that support the findings of this study are available from the corresponding author upon reasonable request.

## Notes

### Competing Interest Statement

The authors have declared no competing interest.

### Author Declarations

The study was approved by the Ethical Review Committee, Institute of Public Health, Lahore, Pakistan, which granted ethical approval for this work. Permission was also obtained from the study setting (Lahore General Hospital, Lahore, Pakistan), and patient consent was obtained before specimen collection and data recording.

